# The Role of AI Model Documentation in Translational Science: A Scoping Review

**DOI:** 10.1101/2023.01.21.23284858

**Authors:** Tracey A. Brereton, Momin Malik, Mark A. Lifson, Jason D. Greenwood, Kevin J. Peterson, Shauna M. Overgaard

## Abstract

**Background:** Translation of artificial intelligence/machine learning (AI/ML)-based medical modeling software (MMS) into clinical settings requires rigorous evaluation by interdisciplinary teams and across the AI lifecycle. The fragmented nature of available resources to support MMS documentation limits the transparent reporting of scientific evidence to support MMS, creating barriers and impeding the translation of software from code to bedside.

**Objective:** The aim of this paper is to scope AI/ML-based MMS documentation practices and define the role of documentation in facilitating safe and ethical MMS translation into clinical workflows.

**Methods:** A scoping review was conducted in accordance with PRISMA (Preferred Reporting Items for Systematic Reviews and Meta-Analyses) guidelines. MEDLINE (PubMed) was searched using MeSH key concepts of AI/ML, ethical considerations, and explainability to identify publications detailing AI/ML-based MMS documentation, in addition to snowball sampling of selected reference lists. To include the possibility of implicit documentation practices not explicitly labeled as such, we did not use “documentation “ as a key concept but rather as an inclusion criterion. A two-stage screening process (title and abstract screening and full-text review) was conducted by an independent reviewer. A data extraction template was utilized to record publication-related information, barriers to developing ethical and explainable MMS, available standards, regulations, frameworks, or governance strategies related to documentation, and recommendations for documentation for papers that met inclusion criteria.

**Results:** Of the total 115 papers, 21 (18%) articles met the requirements for inclusion. Data regarding the current state and challenges of AI/ML-based documentation was synthesized and themes including bias, accountability, governance, and interpretability were identified.

**Conclusions:** Our findings suggest that AI/ML-based MMS documentation practice is siloed across the AI life cycle and there exists a gray area for tracking and reporting of non-regulated MMS. Recommendations from the literature call for proactive evaluation, standards, frameworks, and transparency and traceability requirements to address ethical and explainability barriers, enhance documentation efforts, provide support throughout the AI lifecycle, and promote translation of MMS. If prioritized across multidisciplinary teams and across the AI lifecycle, AI/ML-based MMS documentation may serve as a method of coordinated communication and reporting toward resolution of AI translation barriers related to bias, accountability, governance, and interpretability.

## Introduction

### Background

Artificial intelligence/machine learning-based tools have the potential to revolutionize healthcare with innovative, efficient, and intuitive approaches to care. We refer to these tools broadly as *AI/ML-based medical modeling software* (AI/ML-based MMS), as our core concern is with models in medical care based on the AI/ML approach of algorithmic modeling [1] that finds optimal (often non-causal) correlations rather than traditional statistical theory-based models that try to capture causal processes and underlying mechanisms. This style of modeling introduces novel questions around validation, methodology, communication, coordination, and ethics. We introduce a new term to focus on these specific issues because, while AI/ML-based MMS may be used for clinical decision support (CDS) or be implemented in software as a medical device (SaMD), CDS includes systems that are not AI/ML-based, and the software in SaMD may not include any AI/ML modeling component.

In healthcare, the use of AI/ML-based MMS is expected to lead to high-quality, safe, and effective outcomes for patients. For this to be achieved, an interdisciplinary evaluation process must be conducted throughout the software lifecycle, requiring perspectives from various subject matter experts to create, develop, and integrate the solution. To ensure success, a process must be in place to capture and communicate critical information about the software, such as its intended use, users, decision-making logic, expected performance, and clinical considerations. However, the lack of standardized procedures or resources to capture AI/ML-based MMS functionality limits communication between experts and users, creating implementation barriers that can delay or prevent a project from being deployed. To improve this, standardized resources should be developed to better enable the translation of AI/ML-based MMS into clinical settings.

### Prior Work

An initial literature review was conducted to understand the current state of documentation and impact on the translation of AI/ML-based MMS into clinical practice. Papers obtained through keyword search in PubMed were analyzed and synthesized. The search focused on characterizing the extent of existing materials rather than exploring any potential issues that may have been overlooked. We found no consensus on ‘best practices ‘ for documentation around what we identified as AI/ML-based MMS, however, we recorded relevant available reporting guidelines offered by government and oversight bodies, as well as ethical principles and theoretical guiding frameworks. Overlapping principles prioritized explainability, transparency, accountability, and trustworthiness, but descriptions were found to be highly variable throughout the field [2-4]. Despite its potential, the adoption of AI/ML-based MMS remained fragmented, and there were reports of bias after its deployment that put patient safety at risk, providing inaccurate or skewed outcomes and recommendations, propagating inequalities, and introducing group harm [5-8]. Findings were presented to a group of multidisciplinary stakeholders across the AI/ML life cycle. This workshop highlighted the relevance and urgency for continuing research regarding the current state and challenges of documentation to support the development and translation of AI/ML-based MMS.

## Methods

### Study Design

The findings from the initial literature review and internal research motivated this scoping review to further evaluate the current state and direction of AI/ML-based MMS documentation. The objective of this study was to scope AI/ML-based MMS documentation practices and define the role of documentation in facilitating safe and ethical MMS translation into clinical workflows. Covidence [9], a screening and data extraction software platform developed in accordance with PRISMA (Preferred Reporting Items for Systematic Reviews and Meta-Analyses) guidelines [10], was leveraged to ensure compliance with scoping review standards and facilitate a systematic process to define eligibility criteria, search the literature, screen results, and select evidence for inclusion, and conduct data extraction.

### Define Eligibility Criteria

Key concepts were identified as *artificial intelligence/machine learning, ethical considerations*, and *explainability* to broadly search the literature for evidence of recommendations to support AI/ML-based MMS documentation practices, scope the development of ethical and explainable software, and define the role of documentation for safe and ethical translation. The utilization of broad concepts aimed to account for the range of AI/ML-based MMS, as well as the available recommendations for documentation practices. Keywords and Medical Subject Headings (MeSH) terms were used to support and generate the search query. The identified eligibility criteria were used to generate the search, with limitations including literature found only within the PubMed database; publications dated after 2015; and in journals identified to be relevant to the study objectives as defined by the “find journals “ functionality of the website Jane [11], a website that mines documents in PubMed to find the best matching journals, authors, or articles. All components of the eligibility criteria and search constraints were included in the search query within PubMed, combined through the AND query function, as shown in <u>Multimedia Appendix 1</u>.

### Search and Screen the Literature

Publications were retrieved and imported into Covidence to conduct a two-stage screening process, including title and abstract screening followed by full-text review, completed by an independent reviewer. Papers were excluded if they did not meet the predefined eligibility criteria, as well as those that were not healthcare related or did not evaluate the current state and direction of AI/ML-based MMS documentation and translation.

### Data Extraction

Included papers were analyzed to consolidate evidence of present documentation practices, scope the development of ethical and explainable software, and define the role of documentation in facilitating translation. A data extraction template (<u>Multimedia Appendix 2</u>) was developed and utilized by an independent reviewer within Covidence to define extraction criteria and synthesize data of included papers in a consistent format. To effectively consolidate evidence of current state and challenges to support the objectives of this study, literature was synthesized by bias, accountability, governance, and interpretability, as well detail communicated recommendations.

## Results

### Characteristics of Included Literature

The initial search within PubMed retrieved 115 papers, which were imported into Covidence for screening and extraction, as shown in the PRISMA flow diagram within <u>Multimedia Appendix 3</u>. Title and abstract screening were conducted, and 34 papers (34/115) met the eligibility criteria for inclusion. After title and abstract screening, papers were subjected to full-text reviews where 21 papers (21/34) met the eligibility criteria for inclusion. Of the 21 papers, all included “AI “, “ML “, or “CDS “ within the keywords, 71% included “ethics “ or “bias “ as keywords, and 57% included “explainability “, “interpretability “, “translation “, “governance “, or “policymaking “ as keywords. sRelevance was of importance as the current state was being evaluated, therefore analysis of publication year was conducted; 3 of the papers were published in 2019, 9 in 2020, 7 in 2021, and 2 in 2022. Additional characteristics of the included publications can be found within <u>Multimedia Appendix 4</u>.

Snowball sampling [12] was used to collect materials like documents from governmental and regulatory agencies not indexed in PubMed, referenced in the retrieved documents but not explained with sufficient details or with minimal context. This allowed full summary of existing frameworks, principles, and standards for ethics (<u>Multimedia Appendix 5)</u> and explainability (<u>Multimedia Appendix 6)</u> on which existing literature relies but falls outside of peer-reviewed, indexed literature.

### Current State and Challenges

Existing work focuses on how the translation of AI/ML-based MMS into patient care settings requires collaboration throughout the AI lifecycle and across interdisciplinary specialties of a product team. Further, effective communication and documentation strategies enable collaborative teams to a) report on developments, b) inform system progression, and c) ensure the cross-functional evaluation and documentation of functions from system functionality to ethical considerations [13]. Product teams face challenges involving insufficient collaboration, lack of knowledge continuity, timing of interventions, and deficient or unavailable reporting resources needed to progress production, therefore disrupting translation into clinical practice [14, 15]. Insufficient collaborations are derived from isolated and fragmented development and documentation processes that fail to prioritize knowledge continuity or obtain a broad range of subject matter expertise [14, 16].

The absence of effective tools and best practices to guide translational practices throughout the AI lifecycle contributes to models performing systematically less well on identifiable patient subgroups, and specifically, marginalized subgroups, which is often left undetected at deployment and identified in ex-post reviews, therefore having strong implications on patient safety, tool performance, and the time and cost of corrections [14, 15, 17, 18]. In the U.S., for products that do not fall under the scope of FDA regulation, reporting guidelines (TRIPOD, CONSORT, IEEE, STROBE) exist but are not widely adopted and do not show evidence of driving analysis and documentation throughout all areas of product development and evaluation [19-22]. Guidance for ethics and explainability is expected to meet the needs of a broad range of stakeholders but what constitutes AI ethics and explainability best practices, who determines them, and how principles are applied and regulated in practice remains undefined [14]. Reddy et al. noted that “there is little dialogue or recommendations as to how to practically address these concerns in healthcare, “ suggesting the need for standardized or mandated regulations to guide processes of collaboration, documentation, and compliance, and enhance translational processes with sufficient evidence to support production [14, 20].

Considerations pertaining to ethics and explainability were investigated within the context of AI/ML-based MMS documentation and translation. Current challenges involving bias, accountability, governance, and interpretability were notable themes requiring thorough review.

### Bias

Differential model performance is a limiting factor to the applicability and deployment of AI/ML-based MMS due to the direct implications biased algorithms have on equity and accuracy of use cases [21]. Reddy et al. used the adage “biases in, biases out “ to simplify the influence of bias and how algorithmic decisions and output directly reflect input [20, 22]. When efforts are not taken to increase data diversity and quality, the existing digital divide, social inequalities, and healthcare disparities are widened, penalize underrepresented and at-risk populations, exacerbate vulnerabilities and increase harm [21, 23, 24].. Fragmented processes from which MMS are derived, often from single institutions with single institution data for training, testing, and validation, may incorrectly assume the target population mimics the training data population (demographically, socioeconomically, medically, etc.). This, in turn, limits external reproducibility due to generalizability concerns across populations [25]. If created from single institution data and not externally validated, risks extend beyond appropriate use cases and further jeopardize patient safety and provide insufficient outputs [20, 21, 24-26]. Kerasidou [27] pointed to the regularity of models deployed with inequalities and emphasized the need for evidence regarding prospective evaluation and representativeness, relative to training and deployment environments, to minimize consequences of patient harm and increase model performance, accuracy, fairness, and trust [23].

### Accountability

The introduction of AI solutions into clinical workflows has raised questions regarding accountability. Does a clinician ‘s responsibility extend to decisions made by algorithms, including any harm or poor outcomes [14, 21, 27, 28]? Must clinicians disclose the level of autonomy or receive consent from patients regarding the use of MMS in their care [16, 20-22]? If a clinician ‘s decision contradicts the AI output, how must a clinician proceed and who is liable [15, 20, 26, 28]? Navigating accountability in relation to the intent of MMS requires the constructs of safety, responsibility, autonomy, consent, and trust to be deeply considered throughout the AI lifecycle [15].

With the application and expectation of MMS aiming to enhance clinician decisions and patient outcomes, proactive solutions to navigate responsibility in response to incorrectness or harm is also imperative. While the level of autonomy upon which MMS operates varies, when systems fail, there is an expectation of accountability [20, 21].Contradicting opinions between clinicians and tools present unique problems. There exist different approaches to defensive medicine within AI. For instance, when a clinician and algorithm have contradicting diagnostic decisions, some clinicians are compelled to follow their instinct and practical experience while others are compelled to defer to an algorithm ‘s logic. Regardless of clinical outcome, patients often remain uninformed as to whether the final decision heeds or disregards an AI recommendation [16].

Responsibility and liability were found to be associated with trust of MMS within patient-clinician relationships, as well as public support of the integration of MMS [15, 20, 23, 27]. Concerns surrounding the dehumanization of healthcare are often present when MMS are deployed, such as the risk of clinician over-reliance, disruptions in patient-clinician relationships, management of false societal expectations of AI [16, 21, 27]. Over-reliance reduces or eliminates patient encounters with clinicians, changing the landscape of patient involvement and potentially missing data that otherwise would be collected outside electronic health records (EHRs) [16, 20]. Grote and Berens stated that “facilitating successful collaboration between ML algorithms and clinicians is proving to be a recalcitrant problem that may exacerbate ethical issues in clinical medicine.

### Governance

A gap between ethics and regulation is said to exist with the range of AI/ML-based MMS classifications [20, 23]. While there are a plethora of international standards and guidance documents for building high-quality, safe, and effective medical device software, there is no explicit definition of ethical criteria; however, the impact of the regulatory process does account for many of the identified ethical considerations. This can be partially explained by the lack of standardized ethical principles and reporting requirements [18, 21, 26]. Software developers communicate a desire for increased guidance, resources, and reporting guidelines to develop and deploy ethically designed MMS [14, 28]. While adequate information or recommendations, privacy protection, data integrity, and regulation frameworks may be facilitated by government oversight, evidence suggests AI/ML-based MMS are not deployed with sufficient levels of documentation to support explainability [23]. Without such oversight and resources allocated to MMS, institutions risk inconclusive, inscrutable, and misguided evidence, unfair outcomes, a lack of traceability and transformative effectiveness [26, 28]. Subjecting researchers to self-monitoring of ethical conduct and determining the level of explainability to accompany MMS is of significant concern but may be combatted with increased reporting rigor and guardrails for evaluation and reporting of evidence [4, 14, 18, 20, 23].

While regulators provide guidance on how to deploy safe and effective medical device software, including requiring robust clinical validations and post-deployment monitoring and surveillance programs, these processes may be too burdensome for non-medical AI/ML-based MMS. Questions like “how much accuracy is sufficient for deployment? “, “what level of transparency is required? “, and “do we understand when the model outputs are likely to be unreliable and therefore should not be trusted? “ have made defining expectations and requirements for governance difficult, especially for varying levels of system autonomy and risk [4, 16, 20, 25, 29]. Although frameworks and principles have been developed by various organizations with goals of increasing explainability (FDA, EU GDPR, WHO, IEEE, FAIR), no recognition of established best practices were identified, only recommended principles to abide by, such as those defined within the AI Ethics Principles [15, 24-26, 29, 30]. Reported consequences of fragmented documentation resources and reporting requirements include a lack of interpretability, accountability, validation, transparency, and trust [14, 20-22].

Clinicians, one of the primary end users of MMS, communicate need for documentation resources to serve as training material for intended use, and a means of communicating the objective, functionality, and limitations of MMS to ensure the appropriate and actionable translation of software into practice [14, 16]. Developing explanatory models that satisfy requirements of providing supporting information for clinical decision making proves to be challenging being that clinical decisions are made based on different modalities and reasoning strategies [14, 16]. The complex nature of clinical decisions is said to therefore call for the transparent traceability of the logic leading to output and how clinicians interacted with what they interpreted, therefore requiring guidance as to what constitutes satisfactory explanations of decisions and how such resources should be documented [14, 16, 18, 26].

### Interpretability

While the field points to explainability to facilitate successful documentation and translation, the literature also acknowledges a gap in resources and best practices to achieve interpretable AI/ML-based MMS [4, 14, 30, 31]. Interpretability, defined as understandable explanations of machine learning model outcomes, accounts for how users should be able to understand algorithmic logic and appropriately implement within clinical workflows [4, 17, 32]. Establishing interpretability is functionally difficult without supplemental documentation, creating challenges in understanding decision logic, deploying transparent tools, defining accountability and responsibility (especially with requirements varying with different risk classifications), and threatening patient safety and trust [4, 16, 17, 20, 31].

The complex nature of AI/ML-based MMS, comprising of multifaceted computations that drive decision aiding output, creates challenges in interpretability and initiates debate regarding the prioritization of performance versus interpretability in system development [4, 20]. “Black boxes “ have been found to make clinical application and decision procedures “notoriously hard to interpret and explain in detail, “ limiting the ability to identify and document technical and logical justifications for decisions and conflicting with core values of patient consent and awareness of the role AI in their care [4, 20, 26, 30, 32]. Kerasidou [27] stated that with black-box systems, “the ‘thinking process ‘ by which outcomes are produced is not obvious to those who use the AI, or even to those who develop it “, raising interpretability, explainability, transparency, and justification concerns from developers to clinicians, as well as from clinicians to patients [4, 26, 33]. Amann et al [4] questioned if, due to their complexity, black boxes are even documentable. Without a way to document and report on explanations or interpretations of AI, it is “hard to determine if differences in diagnoses reflect diagnostically relevant differences between patients or if they are instances of bias or diagnostic errors and over/under-diagnosis, “ further emphasizing the need to mitigate and address biases prior to deployment [30]. Once clinicians are no longer able to fully comprehend decisions, they are not able to explain to the patient how certain outcomes or recommendations were derived, impacting patient safety and trust and affecting care plans [4, 17]. The literature describes the necessity to interpret MMS logic because omitting it has been found to “pose a threat to core ethical values in medicine and may have detrimental consequences for individual and public health, “ including evidence of disregard to ethical and regulatory practices, unsuitable clinical application, and making it impossible to investigate and rectify causes of errors; but as Yoon et al argue, the extent to which interpretability is required still needs to be determined [4, 17, 27, 32].

Beyond an understanding of system complexity, interpretability of system functionality was found to be critical for developers, stakeholders, clinicians, and patients to understand a system in relation to clinical applicability and intended use despite varying perspectives and level of AI intellect [19, 32]. The context of appropriate clinical application is especially important to disclose due to the impact of training data on system performance and deployment environment [29]. When explanations see through, analyze, and assess artifacts of MMS throughout design, development, and implementation, the field may anticipate more informed and trustworthy adoption [16, 20, 31].

## Discussion

To address ethical and explainability barriers, enhance documentation efforts, support MMS throughout the AI lifecycle, and promote the translation of MMS, recommendations identified within literature call for proactive evaluation, multidisciplinary collaboration, investigation and validation protocols, guiding standards and frameworks, and transparency and traceability requirements. Such resources are expected to be derived through scientific evidence, reflect clinical practice needs, complement governance regulations like those outlined by the FDA, provide guidance throughout the AI lifecycle, and promote transparency, explainability, accountability, and trustworthiness.

Documentation serving to proactively outline and guide translational processes, as well as encourage multidisciplinary collaboration, are recommended to support system development throughout the AI lifecycle that promotes patient safety, supports appropriate clinical use, and ensures essential testing and validation processes are completed prior to deployment [14, 15, 21, 25] Although a consensus for universal best practices were not recognized in literature, the importance of prioritizing and planning for AI translation from the start of product design with interdisciplinary collaboration was consistently identified [14, 15]. Proactively accounting for translation reflects ex-ante (as opposed to an ex-post) regulation that is “pre-emptive of foreseeable risks and has a more open and participatory character [20, 26]. This approach also promotes collaboration through participatory design from interdisciplinary teams, gaining perspective to identify foreseeable issues within data or use cases and to expose concerns associated with bias and ethics if present [20, 29]. To be effectively proactive and facilitate successful translation, Wiens et al [29] suggest the need for a roadmap for deploying AI/ML-based tools that consists of a stepwise framework with engaged stakeholders to guide production from the beginning (problem formulation) to end (widespread deployment). Within this roadmap, the engagement of stakeholders occurs early in the process to determine clinical relevance, identify appropriate data and collaborators, consider ethical implications and engage with ethicists, rigorous evaluations and reporting on predictions and algorithm code, organizing clinical trials and safety monitorization strategies, and market deployment approaches, all of which ought to be documented and transparent to the interdisciplinary team throughout the product ‘s lifecycle [29, 34].

Explanations, referred to as a “cardinal responsibility of medical practitioners, “ function as an additional safeguard to ensure the reliability of a system ‘s reasoning process, while “trust “ in clinical decisions correlate with the construct of interpretability at all levels of expertise [16, 17]. As such, provider explanations are said to mitigate concerns and threats of overreliance on MMS through the ability to communicate and document the training for decision logic, system output, and role definition in final decisions made for patients [4, 16]. Interpretability to support decision making is especially important for instances of uncertainty and conflict between AI and clinicians to bolster traceability for liability and legal obligations, as well as mitigate ethical concerns [4, 16, 20]. The complexity of reasoning underlying explanations highlights the need to address the varying definitions of explainability, as well as interpretability, and the extent to which they are required to effectively support MMS, as well as addressing the inconsistency and vagueness surrounding bias and the implications on equity and patient safety [16, 17]. These requirements span beyond the scope of one team within a project, making explainability a necessity to be adopted and prioritized from a multidisciplinary perspective in a way that promotes knowledge continuity and interpretability of MMS logic for both technical and translational requirements [4, 14, 17]. Additionally, to ensure the successful design and development of an AI/ML-based MMS, it is important to capture all requirements and risks, including those related to accountability, to define and remain within the defined intended use, establish the allowable autonomy of AI, identify any responsibilities of clinicians and human-AI interactions, and provide evidence for MMS traceability [16, 19-21, 23, 25-28].. To better facilitate successful translation, new software tools can potentially help address these challenges, such as a centralized platform to track and assess the development of AI/ML-based MMS tools throughout their lifecycle; to capture intended use and users; to identify any device risks or potential ethical implications; to document all progress and decisions; and to communicate and share best practices across a central portfolio of tools. Available and proposed standards, guidelines, frameworks, and governance structures to promote ethical and explainable AI/ML-based MMS documentation and translation mentioned within the literature are presented in <u>Multimedia Appendix 5</u> (ethics) and <u>Multimedia Appendix 6</u> (explainability).

### Limitations

Limitations of this study include the restriction of the search to only one database, PubMed, to conduct the scoping review. This limitation likely restricted the available and proposed governance standards, guidelines, and frameworks found. Notably, additional resources found in our previous research were not explicitly mentioned in the literature (e.g., Google ‘s Model Card for model reporting [35], Model Facts Labels [36], DECIDE-AI [37], and AI Factsheets [38]). Therefore, continuing this investigation as other resources are made available (e.g., Coalition for Health AI guardrails, NIST recommendations) is critical. Additionally, due to the exploratory nature of scoping reviews, this study aimed to assess the available academic literature, organize findings within themes and highlight present gaps regarding AI/ML-based MMS documentation and translation practices, rather than research a more defined research question.

### Implications for Practice and Future Development

As indicated by the findings in this scoping review, while there are existing standards, guidelines, frameworks, and governance structures to guide the documentation of AI/ML-based MMS, there is a need for additional resources to provide appropriate guidance related to ethics and explainability, promote safety and efficacy, provide support throughout the AI lifecycle, and reduce present barriers to translation. The available but fragmented and phase specific resources create an opportunity to either streamline and merge complementary standards, guidelines, frameworks, and governance structures, or encourage the development of new resources. Recommendations highlighted from literature may serve to help inform the development of such new resources that provide actionable items to support MMS, in addition to ask questions such as “how do I know if AI is inappropriate? “, “how do I identify potential ethical issues? “, “what do I check to decide if the data are adequate? “, “how do I prepare data and evaluate models? “, “what models do I use and how do I explain my models? “, “how do I audit? “, and “what else do I need to worry about in development and testing? “ to further help create actionable resources.

A possible solution to this issue is to develop a unified model document that creates a framework for easy discovery of the key requirements of an AI-ML-based MMS while allowing for future expandability/customization to fit the needs of each specialty area and possible future requirements. This unified model document could be shared across all levels of model development, from the ideate phase to post deployment monitoring. The document would not only be a way to capture the fundamental features of any AI/ML-based MMS but would provide guidance on how to address issues. For example, if an MMS developer answers a question of explainability as a “black box “ it could provide some basic recommendations of how to expand explainability. The document could be created and translated to a standard mark-up language like XML/JSON. This would allow it to have a common structure for the documentation but allow for consumption by others for implementation (e.g., Epic, Cerner, etc.). Moreover, large institutions that routinely utilize these tools in clinical practice (e.g., healthcare centers) could adopt a risk-based approach, creating a centralized team of experts to standardize the categorization of all AI-ML-based MMS tools and determine the type of control measures, evidence, and overall rigor required for their safe and effective development, deployment, and monitoring.

In addition to the unified document, a possible direction of the industry is to follow others who develop tools and architectures that follow a DevOps framework for ML called MLOps. The first step could be at a national level, either through regulatory bodies (FDA, IEEE, etc.), medical specialties (ACOG, AAFP, ACS, etc.), or through industry and academic collaboration (Google, Amazon, Epic, etc.), to develop a set of practice standards, similar to other regulations such as SaMD, that are adopted across the AI/ML-based MMS industry and require certain documentation and disclosure of MMS technologies that create consistency throughout development to the release to the public. Additionally, organizations could create AI/ML-based MMS translational boards that are comprised of various experts in the fields of AI/ML, including data science/engineering, IT, ethics, EHR, nursing/clinical/translational/pharmacy informatics, and clinical expertise. Like an IRB, found at any research institution, the AI/ML translational board would help address gaps in ethics/bias, explainability, and efficacy and reduce the translational barriers. Lastly institutions and medical colleges could require training and periodic refresher courses for clinicians on AI/ML-based MMS tools that would follow up to date standards of practice with many other medical devices (CLIA, LARC training, BLS/ACLS, etc.).

### Conclusions

The adoption of AI/ML-based MMS tools has “the power to impact the lives of so many, “ therefore calling for comprehensive evaluations that prioritize ethical considerations and the ability to provide explanations that transparently communicate how a decision is reached, what the intended use is, and interpretable by multidisciplinary perspectives, from experts to non-experts. The ability of MMS to deliver on expectations is dependent on resolving barriers to translation, including those related to ethical considerations and explainability. To navigate the potential challenges of trade-offs between diagnostic performance and lack of interpretability, as well as provide traceability to demonstrate compliance, ethics and explainability must be accounted for leveraging available and developing reporting guidelines, standards, frameworks, and governance regulations and oversight. Documentation developed to address identified challenges related to bias, accountability, governance, translation, and interpretability is expected to assist multidisciplinary teams throughout the AI lifecycle and promote necessary evaluation and validation requirements, transparency, trust, standards and frameworks, proactive evaluation, interpretability, and multidisciplinary collaboration.

## Supporting information

Multimedia Appendix 1

Multimedia Appendix 2

Multimedia Appendix 3

Multimedia Appendix 4

Multimedia Appendix 5

Multimedia Appendix 6

## Data Availability

All data produced in the present study are available upon reasonable request to the authors

## Multimedia Appendix

**Multimedia Appendix 1**

Search query (key concepts and search limitations).

[<u>Multimedia_Appendix_1</u>]

**Multimedia Appendix 2**

Data extraction template.

[<u>Multimedia_Appendix_2</u>]

**Multimedia Appendix 3**

PRISMA diagram.

<u>Multimedia_Appendix_3</u>]

**Multimedia Appendix 4**

Included paper characteristics.

[<u>Multimedia_Appendix_4</u>]

**Multimedia Appendix 5**

Available and proposed governance for ethics.

[<u>Multimedia_Appendix_5</u>]

**Multimedia Appendix 6**

Available and proposed governance for explainability.

[<u>Multimedia_Appendix_6</u>]

