## Supplementary material for "The Role of AI Model Documentation in Translational Science: A Scoping Review": Multimedia Appendix 1

**Multimedia Appendix 1.** Key concepts and search limitations used to generate the scoping review query, including the keywords and Medical Subject Headings (MeSH) terms and date and journal constraints.

| **Key Concept** | **Keywords** | **MeSH Terms** | **Key Concept Search Query** |
| --- | --- | --- | --- |
| Artificial Intelligence / Machine Learning | Artificial intelligence, machine learning | "Artificial Intelligence"[Mesh] OR "Machine Learning"[Mesh] OR "Decision Support Systems, Clinical"[Mesh] | "artificial intelligence"[Text Word] OR "machine learning"[Text Word] OR "Artificial Intelligence"[Mesh] OR "Machine Learning"[Mesh] OR "Decision Support Systems, Clinical"[Mesh] |
| Ethical Considerations | Explainability, interpretability, governance, documentation, translation, translational informatics | "Clinical Governance"[Mesh] OR "Documentation"[Mesh] OR "Standard of Care"[Mesh] | explainability[Text Word] OR interpretability[Text Word] OR governance[Text Word] OR documentation[Text Word] OR translat*[Text Word] OR “translational informatics”[Text Word] OR "Clinical Governance"[Mesh] OR "Documentation"[Mesh] OR "Standard of Care"[Mesh] |
| Explainability | ethical consideration*, ethic(s), bias(es), algorithmic bias | "Ethical Analysis"[Mesh] OR "Ethics, Clinical"[Mesh] OR "Selection Bias"[Mesh] | ethical consideration*[Text Word] OR ethic*[Text Word] OR bias*[Text Word] OR "algorithm bias"[Text Word] OR "Ethical Analysis"[Mesh] OR "Ethics, Clinical"[Mesh] OR "Selection Bias"[Mesh] |

|  | **Search Limitation** | **Query** |
| --- | --- | --- |
| Database | PubMed | |
| Date | Publication after 2015 | 2015/01/01:2022/2/1[Date - Publication] |
| Journal | Journals identified to be relevant to the study objectives as defined by Jane (cite Jane) | (Nature medicine"[Journal] OR "J Am Med Inform Assoc."[Journal] OR "PLoS One"[jour] OR “BMJ Open”[Journal] OR "Bull World Health Organ"[jour] OR “IEEE Trans Vis Comput Graph”[Journal] OR “Bioethics”[Journal] OR “J Oral Biol Craniofac Res”[Journal] OR “BMC Med Inform Decis Mak”[Journal] OR “JMIR Med Inform”[Journal] OR “J Med Internet Res”[Journal] OR “Stud Health Technol Inform”[Journal] OR “J Am Coll Radiol”[Journal] OR “Yearb Med Inform”[Journal] OR “Artif Intell Med”[Journal] OR “Clin Radiol”[Journal] OR “Comput Biol Med”[Journal] OR “Comput Methods Programs Biomed”[Journal] OR ”Int J Med Inform”[Journal] OR “Sci Eng Ethics”[Journal] OR “Implement Sci”[Journal] OR “J Med Ethics”[Journal] OR “Comput Inform Nurs”[Journal]) |

**Full PubMed Search Query**:

("Artificial Intelligence"[Mesh] OR "Machine Learning"[Mesh] OR "Decision Support Systems, Clinical"[Mesh] OR "artificial intelligence"[Text Word] OR "machine learning"[Text Word]) **AND** ("Clinical Governance"[Mesh] OR "Documentation"[Mesh] OR "Standard of Care"[Mesh] OR explainability[Text Word] OR interpretability[Text Word] OR governance[Text Word] OR documentation[Text Word] OR translat*[Text Word] OR “translational informatics”[Text Word]) **AND** ("Ethical Analysis"[Mesh] OR "Ethics, Clinical"[Mesh] OR "Selection Bias"[Mesh] OR ethical consideration*[Text Word] OR ethic*[Text Word] OR bias*[Text Word] OR "algorithm bias"[Text Word]) **AND** (2015/01/01:2022/2/1[Date - Publication]) **AND** (Nature medicine"[Journal] OR "J Am Med Inform Assoc."[Journal] OR "PLoS One"[jour] OR “BMJ Open”[Journal] OR "Bull World Health Organ"[jour] OR “IEEE Trans Vis Comput Graph”[Journal] OR “Bioethics”[Journal] OR “J Oral Biol Craniofac Res”[Journal] OR “BMC Med Inform Decis Mak”[Journal] OR “JMIR Med Inform”[Journal] OR “J Med Internet Res”[Journal] OR “Stud Health Technol Inform”[Journal] OR “J Am Coll Radiol”[Journal] OR “Yearb Med Inform”[Journal] OR “Artif Intell Med”[Journal] OR “Clin Radiol”[Journal] OR “Comput Biol Med”[Journal] OR “Comput Methods Programs Biomed”[Journal] OR ”Int J Med Inform”[Journal] OR “Sci Eng Ethics”[Journal] OR “Implement Sci”[Journal] OR “J Med Ethics”[Journal] OR “Comput Inform Nurs”[Journal])
