## Supplementary material for "The Role of AI Model Documentation in Translational Science: A Scoping Review": Multimedia Appendix 2

**Multimedia Appendix 2.** Data extraction template developed on Covidence to standardize information extracted from studies to satisfy scoping review objectives.

| **Data Extraction Template (developed in Covidence)** | |
| --- | --- |
| **General Information of Publication** | Title |
|  | Authors |
|  | Year |
|  | Country |
| **Publication Characteristics** | Study design |
|  | Topics included (AI/ML, Documentation, Governance, Ethical considerations, Explainability, and/or Translation) |
|  | Objective of publication |
|  | Key words |
| **Research Objective Specific Information** | Present challenges in AI/ML |
|  | Ethical considerations (Identified gaps and barriers / Recommendations) |
|  | Explainability (Identified gaps and barriers / Recommendations) |
|  | Standards, regulations, best practices, or governance strategies mentioned |
|  | Overall recommendations from paper |
|  | Relevant quotes |
|  | Additional relevant information |
