## Supplementary material for "The Role of AI Model Documentation in Translational Science: A Scoping Review": Multimedia Appendix 3

**Multimedia Appendix 3.** PRSIMA flow diagram summarizing the two-stage screening process used to conduct the scoping review. The initial search within PubMed retrieved 115 papers that were imported into Covidence for title and abstract screening and full-text reviews in accordance with PRISMA. 21 papers met all eligibility criteria within the search constraints and were selected for inclusion.

115 studies imported for screening

115 studies screened

34 full-text studies assessed for eligibility

115 studies imported for screening

21 studies included

115 studies imported for screening

0 duplicates removed

81 studies irrelevant

13 studies excluded

- 6 wrong study design

- 4 wrong setting

- 2 wrong intervention

- 1 wrong outcome

.
