## Supplementary material for "The Role of AI Model Documentation in Translational Science: A Scoping Review": Multimedia Appendix 4

**Multimedia Appendix 4.** Overview of publications included in the scoping review (N=21).

| Study author(s) / year published | Country | Key words | Study aims |
| --- | --- | --- | --- |
| Allen et al. (2019) [25]  A road map for translational research on artificial intelligence in medical imaging | United States | NA | Summarizes key priorities for translational research developed at the National Institutes of Health/RSNA/ACR/The Academy Workshop to create a road map to grow an ecosystem of will allow robust collaborations between practicing clinicians and AI researchers to advance foundational and translational research. |
| Amann et al. (2020) [4]  Explainability for artificial intelligence in healthcare: a multidisciplinary perspective | Switzerland | Artificial intelligence, Machine learning, Explainability, Interpretability, Clinical decision support | Assess the role of explainability in medical AI and ethically evaluate what explainability means for the adoption of AI-driven tools into clinical practice. |
| Baig et al. (2020) [28]  Urgent need for developing a framework for the governance of ai in healthcare | Saudi Arabia | Healthcare AI, Data science, Ethics, Governance | Discuss the urgent need to institute a robust governance and compliance framework to ensure the efficacy, safety, privacy, and ethical considerations of the application and integration of AI in the healthcare domain. |
| Loi et al. (2020) [31]  Transparency as design publicity: explaining and justifying inscrutable algorithms | Switzerland | Machine learning, Transparency, Explanations, Justifications, Philosophy of science, Computing methodologies~ Artificial intelligence, Cognitive science, Machine learning, Human-centered computing~ HCI theory, Concepts and models | Argue that transparency of machine learning algorithms, just as explanation, can be defined at different levels of abstraction and propose “design publicity,” an approach that explains and provides evidence for algorithms as an intentional product in a given domain, as a new form of algorithmic transparency. |
| Grote et al. (2021) [16]  How competitors become collaborators— Bridging the gap(s) between machine learning algorithms and clinicians | Germany | Clinical expertise, Defensive medicine, human–computer interaction, Machine learning, Medical diagnosis | Evaluate different epistemic and normative factors that may lead to algorithmic overreliance within clinical decision-making and argue that there is an intriguing dialectic in the collaboration between clinicians and ML algorithms. |
| Ho et al. (2021) [24]  Ensuring trustworthy use of artificial intelligence and big data analytics in health insurance | China/United States/Singapore | NA | Discuss what a robust ethical and regulatory environment might look like for big data analytics in health insurance and describe examples of safeguards and participatory mechanisms that should be established. |
| Ho et al. (2019) [26]  Governance of automated image analysis and artificial intelligence analytics in healthcare | Singapore | NA | Discuss the nature of AI governance in biomedicine, as well as its limitations, and argue that radiologists must assume a more active role in propelling medicine into the digital age. |
| Kerasidou (2021) [27]  Ethics of artificial intelligence in global health: Explainability, algorithmic bias and trust | UK | Artificial intelligence, Global health, Lower-middle-income-countries (LMICS), Explainability, Algorithmic bias, Trust | Argue that the pathway to fair, appropriate, and relevant AI necessitates the development and successful implementation of national and international rules and regulations that define parameters and set boundaries of operation and engagement. |
| Landau et al. (2022) [22]  Developing machine learning-based models to help identify child abuse and neglect: key ethical challenges and recommended solutions | United States | Child abuse and neglect, Phenomenological ethics, Machine learning–based risk models, Pediatric emergency departments, Electronic health record | Discuss and provide recommendations for key ethical issues related to machine learning-based risk models development and evaluation, as well as identify several areas for further policy and research. |
| Liaw et al. (2020) [15]  Ethical use of electronic health record data and artificial intelligence: recommendations of the primary care informatics working group of the international medical informatics association | Australia/UK/Canada/United States | Ethics, clinical; Data accuracy; Information systems; Medical record systems, computerized; Artificial intelligence | Using a literature review and Delphi exercises, identified practical recommendations for the curation of routinely collected health data and artificial intelligence in primary care with a focus on ensuring their ethical use. |
| Makridis et al. (2021) [32]  Ethical applications of artificial intelligence: evidence from health research on veterans | United States | Artificial intelligence; Ethics; Veterans; Health data; Technology; Veterans affairs; Health technology; Data | Explored newly developed principles around trustworthy AI and evaluated their relevance for application at scale to vulnerable groups that are potentially less likely to benefit from technological advances. |
| Mökander et al. (2021) [21]  Ethics‐based auditing of automated decision‐making systems: nature, scope, and limitations | UK | Artificial intelligence, Auditing, Automated decision-making, Ethics, Governance | Discuss the feasibility and efficacy of ethics-based auditing (EBA) as a governance mechanism by providing a theoretical explanation of how EBA can contribute to good governance, propose criteria for how to design and implement EBA, and identifying the conceptual, social, economic, organizational, and institutional constraints with EBA. |
| Morley et al. (2020) [14]  From what to how: an initial review of publicly available ai ethics tools, methods and research to translate principles into practices | UK | Artificial intelligence, Applied ethics, Data governance, Digital ethics, Governance, Ethics of AI, Machine learning | Reviewed and presented research on publicly available AI ethics tools to contribute to closing the gap between principles and practices by constructing a typology that may help developers apply ethics at each stage of the ML development pipeline, and to signal to researchers where further work is needed. |
| Ploug et al. (2020) [30]  The four dimensions of contestable AI diagnostics - A patient-centric approach to explainable AI | Denmark/UK/Norway | AI diagnostics, Contestability, Explainability, Health data, Bias, Performance, Organization of diagnostic labor | Suggest that explainability should be explicated as 'effective contestability’ and, taking a patient-centric approach, argue that patients should be able to contest the diagnoses of AI diagnostic systems through the availability of information regarding the AI system's use of data, the system's potential biases, the system performance, and the division of labor between the system and health care professionals. |
| Reddy et al. (2020) [20]  A governance model for the application of AI in healthcare | Australia | Artificial intelligence, Healthcare, Ethics, Regulation, governance Gramework | Propose a governance model that aims to not only address the ethical and regulatory issues that arise out of the application of AI in health care, but also stimulate further discussion about governance of AI in health care. |
| Roski et al. (2020) [19]  Enhancing trust in AI through industry self- governance | United States | Artificial intelligence/Ethics, Artificial intelligence/Organization and administration, Certification, Accreditation, Policymaking | Highlight recently published literature on AI risks and mitigation strategies that would be relevant to designing, implementing, and promoting self-governance, as well as describe a process for how a diverse group of stakeholders could develop and define standards for promoting trust and AI risk-mitigating practices through greater industry self-governance. |
| Samuel et al. (2020) [18]  Defining ethical standards for the application of digital tools to population health research | England | NA | Propose a process of ethics governance for population health research in higher education institutions that takes the form of review after the research has been completed, with particular focus on the role artificial intelligence algorithms play in augmenting decision-making. |
| Stix (2021) [34]  Actionable principles for artificial intelligence policy: three pathways | Netherlands | Artificial intelligence policy, Actionable principles, Ethics, Ethics of artificial intelligence, Governance of artificial intelligence | Propose a novel framework for the development of 'Actionable Principles for AI' that acknowledges the relevance of AI Ethics Principles and methodological elements to increase their practical implementation in policy processes and includes a case study using elements extracted from the Ethics Guidelines for Trustworthy AI. |
| Wiens et al. (2019) [29]  Do no harm: a roadmap for responsible machine learning for health care | United States | NA | Present a framework, context, and guidelines for accelerating the translation of machine-learning-based interventions in health care, highlighting that successful translation will require a team of engaged stakeholders and a systematic process from beginning (problem formulation) to end (widespread deployment). |
| Yoon et al. (2021) [17]  Machine learning in medicine: should the pursuit of enhanced interpretability be abandoned? | UK/United States | NA | Argue why interpretability should have primacy alongside empiricism, and detail reasonings for the importance of interpretability, such as the establishment of trust and professional and public acceptance. |
| Zhao et al. (2021) [23]  Ethics, integrity, and retributions of digital detection surveillance systems for infectious diseases: systematic literature review | China | Artificial intelligence, Electronic medical records, Ethics, Infectious diseases, Machine learning | Conducted a systematic review to investigate ethical issues identified from utilizing artificial intelligence-augmented surveillance or early warning systems to monitor and detect common or novel infectious disease outbreaks to inform necessary ethical considerations for medical practitioners and policymakers and global governance structures. |
