## Supplementary material for "The Role of AI Model Documentation in Translational Science: A Scoping Review": Multimedia Appendix 5

**Multimedia Appendix 5.** Available and proposed standards, guidelines, frameworks, and governance structures to promote ethical AI/ML-based MMS documentation and translation mentioned throughout literature.

| Available | |
| --- | --- |
| Governance for Ethical Principles | |
| Ethics-Based Auditing (EBA) [21] | - Governance mechanism available for entities to control or influence the design and deployment of automated decision-making systems (ADMS) by providing a structured process whereby behavior can be assessed throughout the product lifecycle (conceptualization, design, deployment, and use of ADMS) for consistency and adherence to predefined ethical principles - EBA aims to contribute to good governance approaches and be an integral component of managing ethical risks of CDS tools, helping to identify, visualize, and communicate ethics within a CDS system, reveal the casual chain behind decisions through improved procedural transparency and regularity method, and allow stakeholders to identify who should be held accountable for potential ethical harms - EBA framework procedure:  1. Describe the purpose of the ADMS 2. Define the standards or verifiable criteria based on which the ADMS should be assessed 3. Disclose the process, including a full account of the data use and parties involved 4. Assess the impact the ADMS has on individuals, communities, and its environment 5. Evaluate whether the benefits and mitigated risks justify the use of ADMS 6. Determine the extent to which the system is reliable, safe, and transparent 7. Document the results and considerations 8. Reflect and evaluate periodically, i.e., create a feedback loop  - *Available EBA frameworks: Framework to operationalize AI (AIEIG), Privacy impact assessment (CNIL), AI impact assessment (ECP), Guidelines for trustworthy AI (EU), AI: Australia’s ethics frameworks (Gov. of Australia), Algorithmic impact assessment (Gov. of Canada), Model AI governance framework (PDPC), AI ethics principles and guidelines (Smart Dubai)* - *Available EBA tools: FAIRVIS (CMU), What-if-tool (Google), AI Fairness 360 (IBM), Fairlean (Microsoft), Turingbox (MIT), Responsible AI toolkit (PwC), Aequitas (University of Chicago), CERTIFAI (University of Texas)* |
| AI Ethics Principles [34] | - Concept developed to condense complex ethical considerations and requirements into a format that can be accessible to both the developers and users of AI technology, therefore aiming to create standards for AI ethics - Function to help organizations by informing and providing guidance on AI ethics, but often fail to be actioned in governmental policy - *Example of AI Ethics Principles: Ethics Guidelines for Trustworthy AI (EU AI HLEG), AI4People, Five Principles Key to Any Ethical Framework for AI, Recommendations of the Council of Artificial Intelligence, Beijing AI Principles for R&D* |
| Ethics Guidelines for Trustworthy AI (EU) [21, 14, 34] | - Prepared by the High-Level Expert Group on Artificial Intelligence (AI HLEG) and provides guidance on the implementation and realization of trustworthy AI through the definition of ethical principles and seven key requirements, forming the basis for legal obligations for high-risk AI system deployment within EU and satisfying the three components that make up Trustworthy AI (lawful, ethical, and robust) that must be met throughout the system’s entire life cycle - Defined key ethical principles as:   - Respect for human autonomy   - Prevention of harm   - Fairness   - Explicability - Defined key requirements as:   - Human agency and oversight   - Technical robustness and safety   - Privacy and data governance   - Transparency   - Diversity, non-discrimination, and fairness   - Societal and environmental wellbeing   - Accountability - Framework was dynamically co-developed through an agile process with stakeholders to support actionability and allow for multidimensional input on key requirements, including assistance of experts from multiple sectors (ethicists, lawyers, machine learning researchers, etc.), engagement with European AI Alliance, and feedback from the public |
| AI4People [14] | - Defined key ethical principles as:   - Beneficence   - Non-maleficence   - Autonomy   - Justice   - Explicability |
| Five Principles Key to Any Ethical Framework for AI [14] | - Defined key ethical principles as:   - AI must be beneficial to humanity   - AI must not infringe on privacy or undermine security   - AI must protect and enhance our autonomy and ability to take decisions and choose between alternatives   - AI must promote prosperity, and AI systems must be understandable and explainable |
| Recommendation of the Council of Artificial Intelligence (OECD) [14] | - Defined key ethical principles as:   - Inclusive growth, sustainable development and well-being   - Robustness, security and safety   - Human-centered values and fairness   - Transparency, explainability, and accountability |
| Beijing AI Principles for R&D (Beijing AI Principles) [14] | - Defined key ethical principles as:   - Do good - covers the need for AI to promote human society and the environment   - Be responsible - covers the need for researchers to be aware of negative impacts and take steps to mitigate them   - Control risks - covers the need for developers to improve the robustness and reliability of systems to ensure data security and AI safety   - For humanity - covers the need for AI to serve humanity by conforming to human values including freedom and autonomy   - Be diverse - covers the need for AI to benefit as many people as possible   - Be ethical - covers the need to make the system as fair as possible, minimizing discrimination and bias and the need for AI to be transparent, explainable, and predictable |
| National Academy of Medicine (NAM) AI Implementation Life Cycle [19] | - The National Academy of Medicine (NAM) outlined an AI implementation life cycle that serves as an organizing schema to understand specific AI risks and mitigation practices - The phases included within the life cycle include:   - Phase 1 – Needs Assessment – Identify or Reassess Needs, Describe Existing Workflows   - Phase 2 – Development – Define the Desired Target State, Acquire or Develop AI System   - Phase 3 – Implementation – Implement AI System in Target Settings   - Phase 4 – Maintenace – Maintain, Update, or De-Implement, Monitor Ongoing Performance |
| Ethically Aligned Design [21, 19] | - The Institute of Electrical and Electronics Engineers (IEEE) published Ethically Aligned Design to provide directional insight and actionable recommendations (legal and technical) through principles, standards, and regulations for technologists, educators, and policy makers - Ethically Aligned Design calls for the ethical and values-based design, development, and implementation to be guided by 8 principles, including: human rights, well-being, data agency, effectiveness, transparency, accountability, awareness of misuse, competence |
| 2016 Privacy, Ethics, and Data  Access Framework for Real World EHR  Data [15] | - Framework developed by the International Medical Interpreters Association (IMIA) that lists 14 ethical principles to guide data custodianship and appropriate access to big data by various stakeholders (incorporates FAIR guiding principles as well for data management): Autonomy, Respect rights and dignity of patients, Respect clinical judgment, Duty to provide care, Protection of the public from harm, Beneficence, Justice, Non-maleficence (obligation to not inflict harm intentionally), Reciprocity, Solidarity, Stewardship, Trust, Lawfulness, and Transparent project approval process |
| Strengthening the Reporting of Observational Studies in Epidemiology (STROBE) Checklist [22] | - The STROBE checklist includes 22 items considered essential for good reporting that provides guidance on how to report observational research within cohort, case-control, and cross-sectional study designs - The checklist aims to improve the quality of reporting of observational studies and promote validation and reproducibility of AI, with defined sections of the checklist including title and abstract, introduction, methods, results, discussion, and other information |
| Transparent Reporting of a Multivariable Prediction Model for Individual Prognosis or Diagnosis (TRIPOD) Checklist [20] | - The TRIPOD checklist includes 22 items considered important for transparent reporting of predictive models including model specification, performance, and validation - The checklist aims to improve and promote transparency when reporting upon AI models, with defined sections of the checklist including title and abstract, introduction, methods, results, discussion, and other information |
| FAIR Initiative [25] | - Initiative to produce datasets that are findable, accessible, interoperable, and reusable (FAIR), functioning to fulfill high-quality goals of reliability, reproducibility, and explainability - The four foundational principles of FAIR serve to guide a range of stakeholders, from data producers to publishers, in interacting with data, algorithms, tools, and workflows for data management and stewardship |
| Proposed | |
| Actionable Principles for AI [34] | - Proposed preliminary framework to support and improve the implementation of AI Ethics Principles in governmental policy, acknowledging the relevance of AI Ethics Principles but promoting methodological elements to increase practical implementability and actionability in policy processes for governmental actors - To be successful, Actionable Principles for AI must develop A) a succinct condensation of broad and deep ethical theories into an accessible number of principles and B) strike a balance between pursuing an ideal hypothetical outcome, and working to secure workable pragmatic outcomes - Follows three propositions from inception to development to post-publications:   1. Preliminary landscape assessment to address the contextual environment within which Actionable Principles arise   2. Multi-stakeholder participation and cross-sectoral feedback   3. Mechanisms to support implementation and operationalizability |
| Governance Model for Artificial Intelligence in Healthcare (GMAIH) [20] | - The proposed Governance Model for Artificial Intelligence in Healthcare (GMAIH) aims to address ethical (bias, privacy, and trust), regulatory, safety, and quality concerns throughout phases of AI development to integration - The four main components of the governance model include:   - Fairness - Data Governance Panels to oversee data collection and use, design of AI models to ensure procedural and distributive justice   - Transparency – transparency of model decision making, support for patient and clinician autonomy   - Trustworthiness – educating clinicians and patients about AI, fully informed consent from patients to use AI, appropriate and authorized use of patient data   - Accountability – regulation and accountability at the approval (including FDA SaMD regulation), introduction, and deployment (including TRIPOD checklist for guidance constituting the reporting framework) phases of AI application |
| Industry Self-Governance [19] | - Governmental presence was proposed through industry self-governance, filling a critical gap to construct a more comprehensive approach to the governance of AI solutions through multi-stakeholder participation, consensus of goals and frameworks to enhance trust and regulate/measure adherence to standards and practices, operationalize program design, establish periodicity for recertification and accreditation, create market demand, and evaluate program effectiveness and adherence to best practices - Self-governance was said to be created by an initial group of AI developers, implementers, and other stakeholders, calling for active and voluntary participation to identify, implement, and monitor adherence to risk mitigation practice standards - Success of self-governance is dependent on stakeholders' confidence in that the standards and verification methods were developed by appropriately balancing perspectives of consumers/patients, clinicians, AI developers, AI users, and others, as well as effectively complementing available legislative and regulatory efforts |
| Model for ethical scrutiny of AI-based research [18] | - Proposed an ex-post review model for ethical scrutiny of AI-based research functioning to provide an extra layer of governance in the ethics ecosystem after research has been conducted, addressing the present ethics gap, combating inconsistency, and igniting a process of developing a new shared understanding of ethics best practice for AI research - Consists of two layers, the first requiring a systematic, open-science infrastructure for centralized national repositories for open-source algorithms and affiliated data, and the second being the ex-post review process to validate research processes and algorithms to safeguard public health and minimizing risks (said to be potentially conducted by committee comprising academic and stakeholders with a range of expertise across disciplines) |
