## Supplementary material for "The Role of AI Model Documentation in Translational Science: A Scoping Review": Multimedia Appendix 6

**Multimedia Appendix 6.** Available and proposed standards, legislation, organizations, guidelines, frameworks, and governance bodies to promote explainable AI/ML-based MMS tool documentation and translation mentioned throughout literature.

| Available | |
| --- | --- |
| Available Governance Organizations | |
| Food and Drug Administration (FDA) [25, 28, 26, 21, 19] | - The U.S. Food and Drug Administration (FDA) serves to protect public health by ensuring the safety, efficacy, and security of a range of health care products through regulatory guidance, including the regulation of medical devices and computer-aided diagnosis software and algorithms that provide decision-making support to medical practitioners - Basic FDA regulatory requirements manufacturers of medical devices distributed within the U.S. must comply with include Establishment Registration, Medical Device Listing, Premarket Notification 510k or Premarket Approval (PMA), Investigational Device Exemption (IDE), Quality System (QS) Regulation, Labeling Requirements, and Medical Device Reporting (MDR) |
| AI High-Level Expert Group (HLEG) [18] | - AI High-Level Expert Group on Artificial Intelligence (AI HLEG) is an independent expert group set up by the European Commission in June 2018 to support the EU’s vision of “(I) increasing public and private investments in AI to boost its uptake, (ii) preparing for socio-economic changes, and (iii) ensuring an appropriate ethical and legal framework to strengthen European values” - The HLEG is mandated with drafting the AI Ethics Guidelines and Policy and Investment Recommendations deliverables |
| Organization for Economic Co-operation and Development (OECD) [24, 21, 14] | - The Organization for Economic Co-operation and Development (OECD) claims to be the first intergovernmental standard on AI (30+ member countries), designed to ensure AI systems are robust, safe, fair, and trustworthy - This international organization works to build better policies for better lives by establishing evidence based international standards together with governments, policy makers, and citizens - The OECD supports governments by measuring and analyzing economic and social impacts of AI technologies and engages with all stakeholders to identify good practices for public policy - The OCED AI Principles to promote the use of AI in an innovative and trustworthy way and respect human rights include:   - Inclusive growth, sustainable development, and well-being   - Human-centered values and fairness   - Transparency and explainability   - Robustness, security, and safety   - Accountability |
| World Health Organization (WHO) [24] | - The World Health Organization (WHO) is the United Nations agency with 194 Member States that functions to connect nations, partners, and people to promote health, safety, and serve the vulnerable - The WHO collaborates with governments, international organizations, foundations, researchers, and health workers and is rooted in basic principles of the right to health and well-being for all people - WHO defined six principles to guide algorithm development:  1. Protect human autonomy 2. Promote human well-being, safety, and public interest 3. Ensure transparency, explainability, and intelligibility 4. Foster responsibility and accountability 5. Ensure inclusiveness and equity 6. Promote AI that is responsive and sustainable |
| Available Frameworks, Standards, Principles | |
| Common Data Elements (CDEs) [25] | - Standardized key terms and concepts functioning to structure data and enhance data quality throughout AI development and deployment, ensuring AI algorithms perform as expected across a variety of settings - A registry of CDEs can be created to integrate appropriate examination data, create outputs that are standardized, and present final interfaces in a consistent form and structure for users |
| US 21^st^ Century Cures Act [17] | - Regarding Health IT, the 21^st^ Century Cures Act was enacted to catalyze medical product development and funding, functioning to advance innovations that improve the quality of care for patients more efficiently - The law helps get more innovative, low-risk (unrelated to diagnosis, prevention, or treatment of disease) technology into patients’ hands and places a strong emphasis on interoperability among EHRs |
| Promoting the Use of Trustworthy AI in the Federal Government [32] | - In December 2020, the U.S Federal Government enacted legislation that established principles of trustworthy use of AI, requiring agencies designing, developing, acquiring, and using AI within the Federal Government to adhere to:   - Lawful and respectful of our Nation’s value   - Purposeful and performance-driven   - Accurate, reliable, and effective   - Safe, secure, and resilient   - Understandable   - Responsible and traceable   - Regulatory monitored   - Transparent   - Accountable |
| Recommendations on Digital Interventions for Health System Strengthening [24] | - Guideline published by WHO to present recommendations based on critical evaluation of the evidence on emerging digital health interventions, aiming to equip health policymakers and other stakeholders with recommendations and implementation considerations for making informed investments - WHO states these guidelines, targeted at decision makers and public health practitioners, promote and support digital health interventions in healthcare and aim to strengthen evidence-based decision-making on digital approaches by governments and partner institutions (encouraging mainstreaming and institutionalization of effective interventions but also acknowledging limitations) - Includes Principles for Digital Development (designing with users, understanding the ecosystem, reuse of and improvement upon existing digital solutions, and addressing privacy and security concerns) to help implementers integrate established best practices into digital interventions, avoid common pitfalls, and encourage the adoption of approaches that have demonstrated value over time |
| Software as a Medical Device (SaMD) Certification Program [25, 28, 26, 21, 19] | - FDA developed harmonized framework and principles for Software as a Medical Device (SaMD) that enables stakeholders, including regulators, to promote safe innovation and protect patient safety through assessment and certification requirements - FDA SaMD Working Group (WG) developed key definitions for SaMD, framework for risk categorization for SaMD, the Quality Management System for SaMD, and the clinical evaluation of SaMD that must be adhered to achieve SaMD compliance - FDA SaMD, although it provides a reference point as a constructive AI governance framework, is narrow and said to only require regulation of a small portion of medical devices deployed in healthcare through the risk-based approach |
| General Data Protection Regulation (GDPR) [24, 26, 15, 30] | - The General Data Protection Regulation (GDPR) is a digital health intervention regulation passed by the EU that promotes trustworthy, ethical, and human-centric AI through a system of laws and requirements enforced through social and governmental institutions - This regulation focuses on data protection and privacy, seeking to secure the autonomy, dignity, and security of people through key regulatory points involving data protection principles, accountability, data security, data protection by design and by default, consent, and privacy rights |
| Proposed | |
| Proposed Frameworks, Principles, Guidelines | |
| Design publicity [31] | - Proposed new form of algorithmic transparency labeled as “design publicity” that functions to explain algorithms as an intentional product, that serves a particular (or multiple) goals in a given domain of applicability, and provides a measure of the extent to which a goal is achieved and evidence about the way the measure has been reached - Design publicity provides information on:  A. The goal the algorithm is designed to pursue and the moral constraints it is designed to respect (value transparency) B. The way this goal is translated into a problem that can be solved by machine learning (translation transparency) C. The performance of the algorithm in addressing problems (performance transparency) D. Proof of the fact that decisions are taken by consistently applying the same algorithm (consistency transparency)  - While post-hoc explanations do not provide useful elements to judge and debate the justification of such systems, design publicity serves to enable discussions involving the justification, use, and design of an algorithm and each individual decision made |
| Artificial Intelligence Development Act (AIDA) [19] | - An Artificial Intelligence Development Act (AIDA) was suggested to task an organization or government agency with certifying the safety of a broad range of AI products and systems across industry sectors - Suggested to be needed because FDA’s SaMD represents only a small portion of AI solutions deployed in health and healthcare, therefore requiring additional legislation to manage a broader range of AI health solutions |
